# The Effects of Decreased Frame Rate During Cardiac Catheterization on Patient and Radiation Safety Outcomes

**DOI:** 10.1101/2023.10.04.23296575

**Authors:** Al-Ameen Oredegbe, Matthew Derakhshesh, Ravi Vuthoori, Mikhail Torosoff, Anthony Nappi, Neil Yager

**Author notes:** Corresponding author: Al-Ameen Oredegbe MBBChBAO, 43 New Scotland Ave, Albany NY 12208.

## Abstract

**Objective:** Coronary artery disease (CAD) is an important cause of morbidity and mortality all over the world. Cardiac catheterization is one of the most important treatment modalities in managing CAD. Unfortunately, cardiac catheterization is associated with significant ionizing radiation exposure to both patients and personnel. This study aimed to compare the safety and efficacy of a lower video frame rate of 7.5-fps compared with the standard frame rate of 15-fps.

**Materials and Methods:** We retrospectively collected and reviewed data from 84 cardiac catheterizations performed between January and February 2020 at a single tertiary center. The patients were divided into two groups based on frame rate: 15-fps (n=42) and 7.5-fps (n=42). We compared the two groups in terms of demographic data, procedural characteristics, radiation dose, and patient outcomes.

**Results:** Cumulative air kerma was significantly lower in the 7.5-fps group (266.576 mGy) compared to the 15-fps group (524.140 mGy), p=0.0018. Similarly, total dose area product was lower in the 7.5-fps group (15335.617 mGy × cm^2^ compared to the 15-fps group (34784.095 mG × cm^2^), p = 0.0003. Despite this, total fluoroscopic time did not differ between the two groups 7.74 minutes in the 15-fps group and 8.462 minutes in the 7.5-fps group, p = 0.4023). In addition, 30-day mortality 0% in both groups and there were no differences in the number of repeat PCIs performed within 30 days (6 in 7.5-fps and 3 in the 15-fps group), p = 0.2899.

**Conclusion:** During cardiac catheterization, a fluoroscopy rate of 7.5-fps is associated with lower radiation dose compared to 15-fps without an associated increase in fluoroscopy time, mortality or repeat PCI. Therefore, interventionalists should consider using a fluoroscopy rate of 7.5-fps during cardiac catheterization.

## Introduction

Coronary artery disease is one of the major causes of morbidity and mortality in the world. Consequently, the use of cardiac catheterization in diagnosis and treatment is more important than it has ever been.^1^ As technological advances are made in coronary angiography, increasingly complex procedures are being performed.^2^ These include interventions on chronic total occlusions and bifurcational percutaneous coronary interventions (PCIs), amongst others.^3^ Furthermore, the transradial approach is increasingly being utilized worldwide for diagnostic and interventional procedures due to reduced bleeding complications, earlier ambulation, and improved patient comfort.^4^ Unfortunately, cardiac catheterization is associated with significant radiation exposure to both patient and cardiac laboratory personnel. Complex procedures tend to be associated with higher amounts of radiation exposure. Additionally, a substudy of RIVAL (Radial Versus Femoral Access for Coronary Intervention) showed that transradial approach coronary angiography causes increased operator and patient radiation exposure in low-volume radial centers.^3^

Therefore, interventional cardiologists are among medical personnel with the highest exposure to ionizing radiation.^5^ During the course of their careers, interventional cardiologists are exposed to an amount of radiation equivalent to about 2500 to 10,000 chest x-rays. The result of this is a cancer risk of about 1 in 100.^5^ The latter is an example of the stochastic effects of radiation; this is where the risk of an outcome is proportional to the cumulative dose received, e.g. malignancy. In contrast, deterministic effects present after a certain threshold of radiation is exceeded, e.g. skin erythema and ulceration.

Several strategies for mitigating radiation exposure have been utilized in the cardiac catheterization laboratory. These include avoiding left anterior oblique (LAO) and steep caudal/cranial angulation, greater distance from the X-ray tube, lead shielding, suspended protection systems, and vascular robotics.^6^ X-ray generation occurs in an evacuated glass envelope containing a cathode filament and an anode.^7^ The cathode filament is heated, and a high voltage is applied across the gap between the cathode and anode causing the cathode to emit electrons that release energy when they strike the anode. When these electrons meet the anode, a portion of the energy carried by the electrons is transformed into X-rays. In most X-ray systems, electrical current is generated in a pulse train mode. This leads to the production of X-ray pulses instead of continuous irradiation. The X-ray beam emanates from the tube and is modulated by exposed tissue, producing an exit beam that is detected by a detector that transmits this information to a digital video processor. The video frame rate must be a whole number multiple of the X-ray pulse rate to preserve synchrony between X-ray pulse and video frame generation. The standard video frame rate is 30 frames/second; therefore, X-ray pulses are typically 7.5, 15, or 30 pulses per second.^7^

There is some data to suggest that utilizing lower frame rate fluoroscopy during cardiac catheterization can further decrease radiation exposure. In this study we aimed to investigate the default frame rate of 15 frames per second (fps) compared to a lower frame rate of 7.5-fps during coronary angiography and percutaneous coronary intervention (PCI).

## Methods

### Study Population

This was a retrospective, single tertiary center, IRB approved cohort study of patients who underwent cardiac catheterization between January and February 2020. No informed consent was required, and procedures followed were in accordance with institutional guidelines. The study cohort consisted of 84 patients, 42 of whom underwent cineangiography with a frame rate of 15-fps while the other 42 were exposed to a frame rate of 7.5 fps. There was a radiation physicist on site who monitored for radiation exposure in the catheterization laboratory and did not make any changes to the cineangiography or fluoroscopy settings during the study period. The inclusion criteria were patients who underwent left heart catheterization (with or without percutaneous coronary intervention). The exclusion criteria were any electrophysiological studies, transvenous pacemaker implantations alone, or any structural procedures such as balloon valvuloplasties, valve implantations, left atrial appendage occlusions and atrial septal defect closures.

### Data

Demographic data including age, gender, body mass index (BMI), smoking history and cardiac risks factors including a history of diabetes, hypertension, hyperlipidemia, and previously defined coronary artery disease were collected to ensure uniformity between the two groups. Primary outcomes include average air kerma as stratified by the number of stents placed and defined in milligray (mGy), total dose area product, post procedure creatinine measured 24 to 48 hours after catheterization and total fluoroscopy time. Secondary outcomes include 30-day repeat PCI and 30-day mortality.

### Statistical Analysis

Statistical analysis was performed using SPSS software. Chi-square tests were used to analyze categorical variables between the two groups which included most patient demographics apart from age and these categorical variables were expressed as frequencies. An analysis of variance (ANOVA) was performed when comparing the ages between the two frame rate groups and they were expressed as means. Similar analyses were utilized to compare the primary and secondary outcomes between the two groups. A p-value of <0.05 was measured as statistically significant.

## Results

### Patient Demographics and Procedure Characteristics

Of the 82 patients in this study, half underwent angiography with a frame rate of 15 fps while the half were exposed to a frame rate of 7.5 fps. The demographics between both groups which included age, gender, history of coronary artery disease, diabetes mellitus, hypertension, hyperlipidemia, and smoking status were similar (Table 1). Patients who ever smoked were subdivided further into current and former smokers with current smokers comprising 33% of the 15-fps group and 22% of the 7.5-fps group (p = 0.111). In terms of the procedure characteristics, the frequency of a radial approach for access was similar between the two groups (74% for the 15-fps group and 75% for the 7.5-fps group, p = 0.801). When considering PCI, there was also no significant difference in the number of stents deployed between both groups (Figure 1).

**Table 1:**
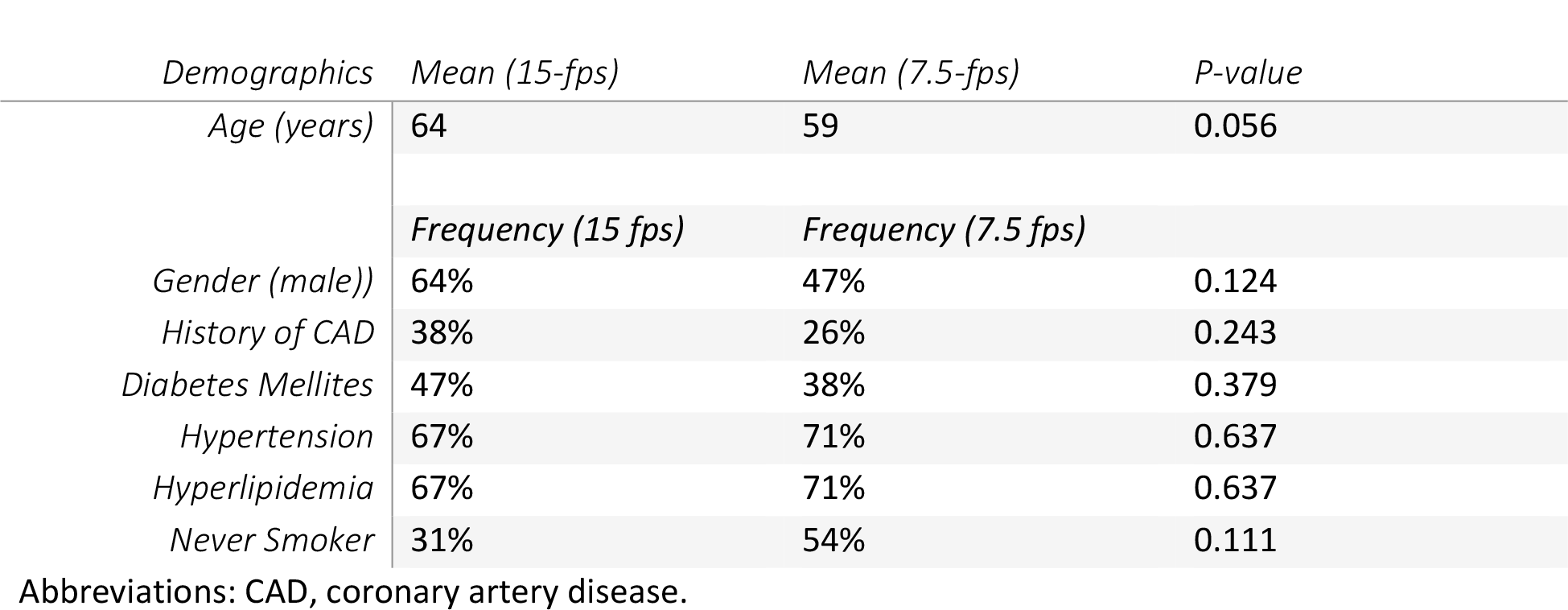
Patient Demographics.

**Figure 1:**
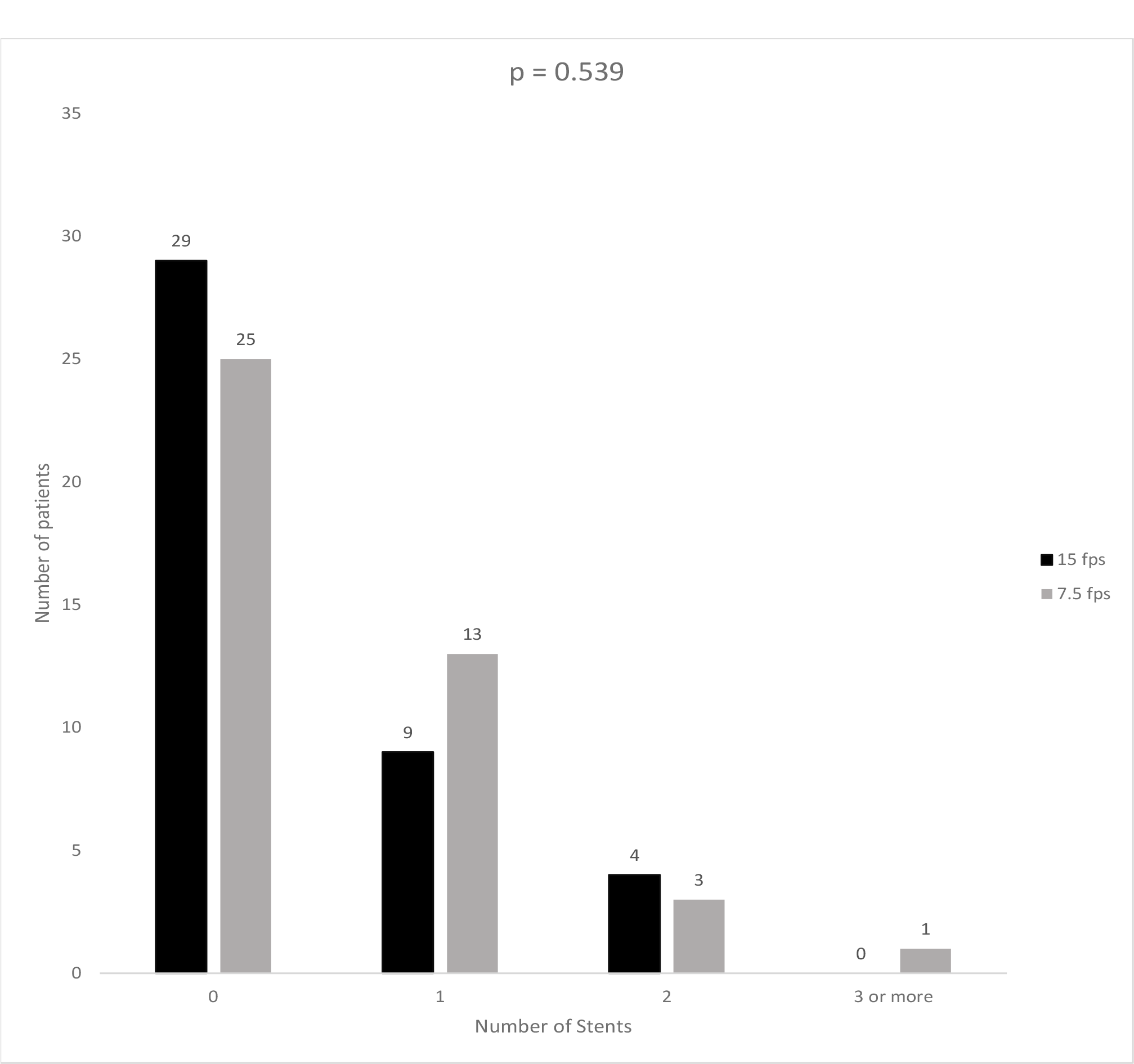
Total number of patients receiving a certain number of stents in the 15-fps and 7.5-fps groups of patients undergoing cardiac catheterization. Black bar: 15-fps group Grey bar: 7.5-fps group

**Figure 2:**
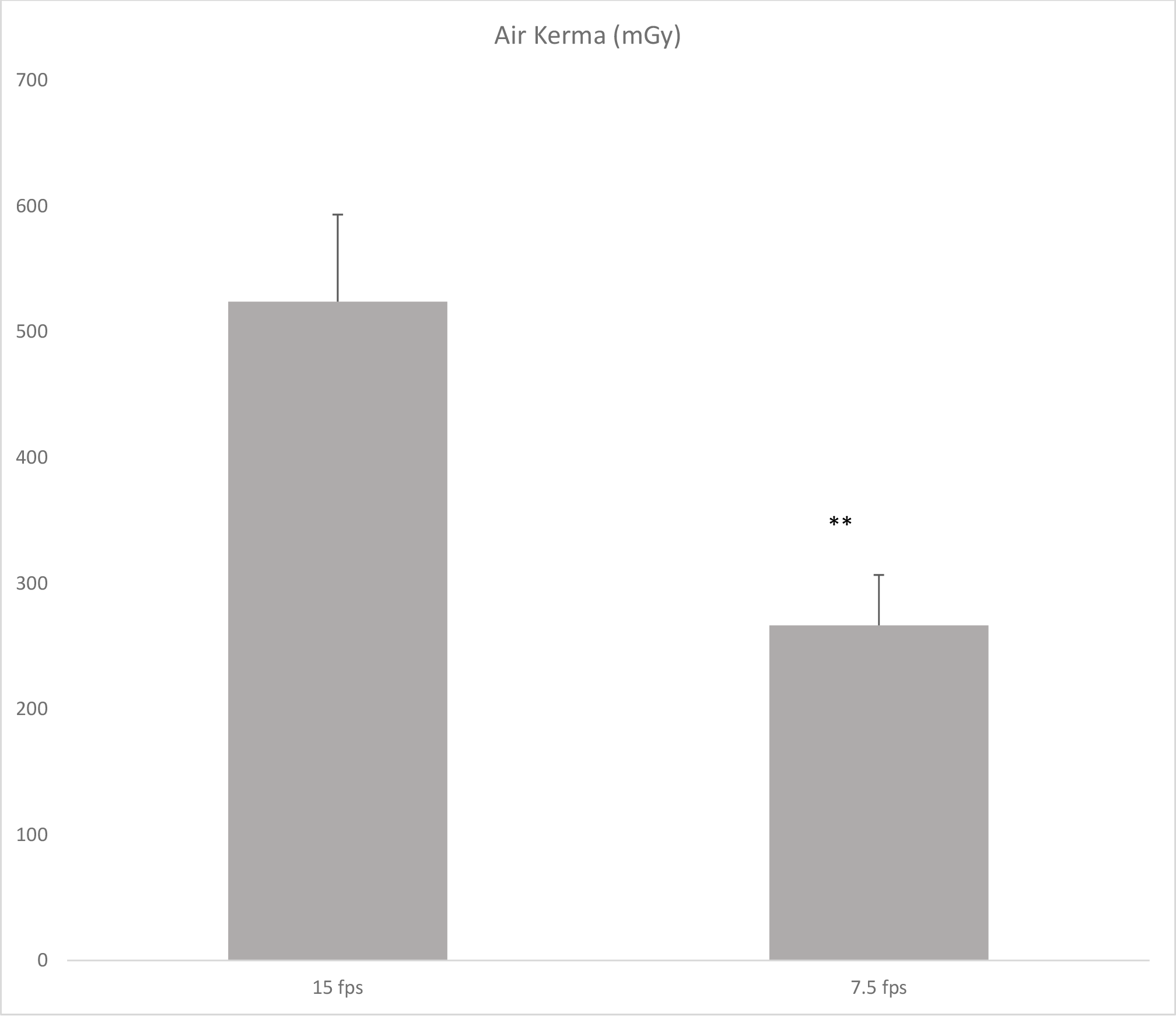
Average air kerma (mGy) measured amongst patients undergoing cardiac catheterization with fluoroscopy rates of 15-fps and 7.5-fps.

**Figure 3:**
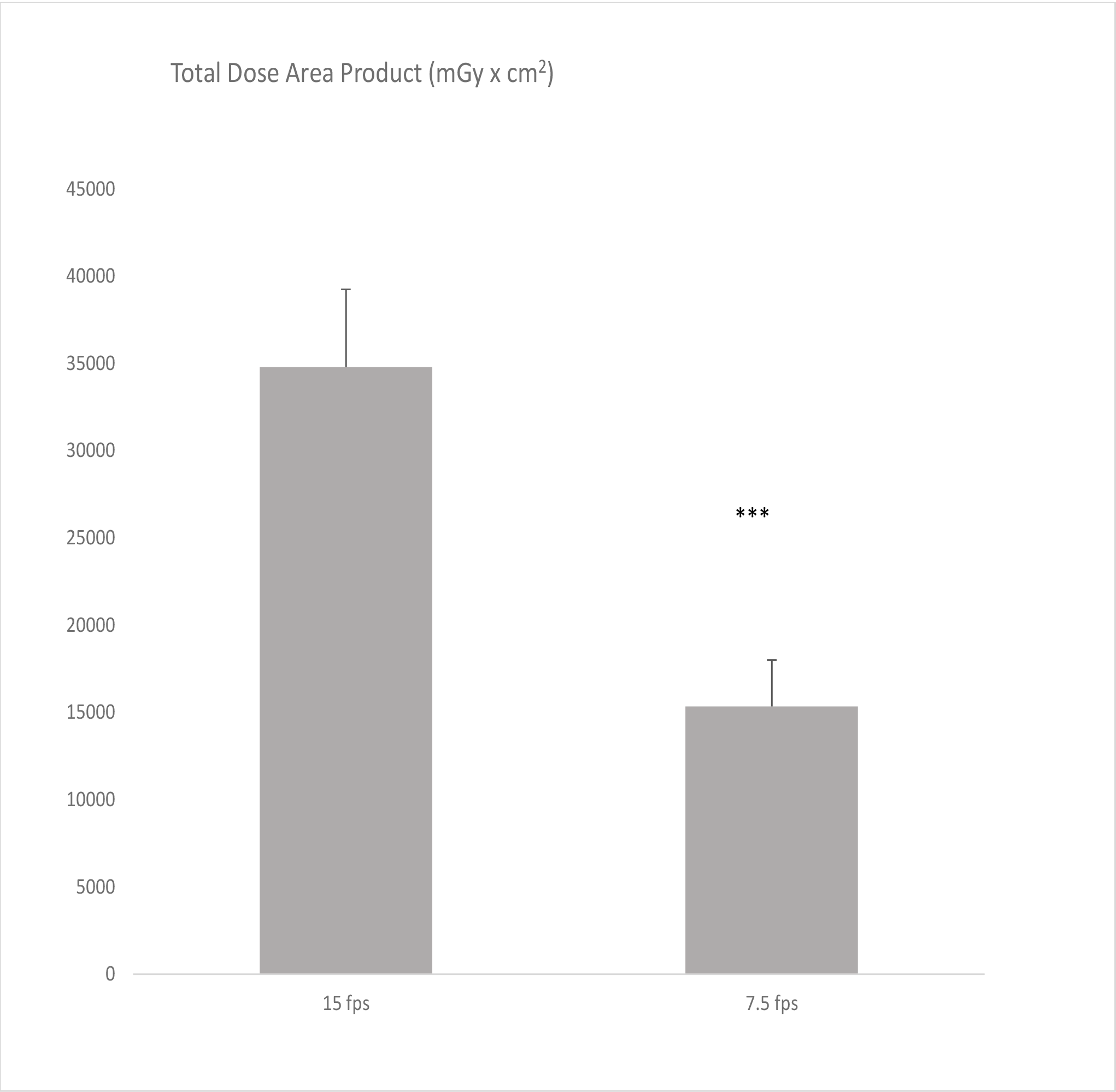
Average total dose area product (mGy × cm^2^) measured amongst patients undergoing cardiac catheterization with fluosocopy rates of 15-fps and 7.5-fps

**Figure 4:**
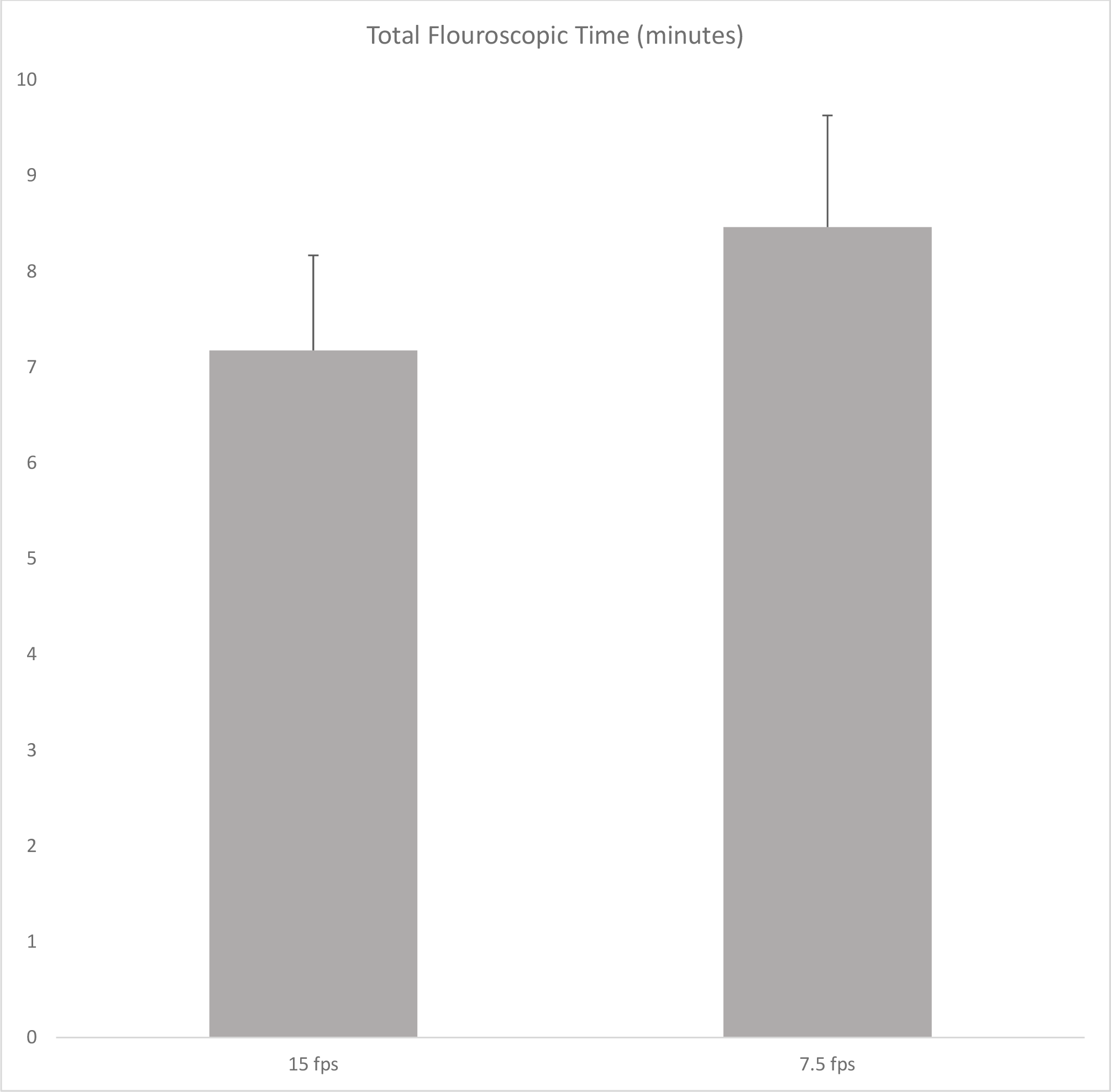
Total fluoroscopy time (minutes) amongst patients undergoing cardiac catheterization at 15-fps and 7.5-fps fluoroscopy rates.

**Figure 5:**
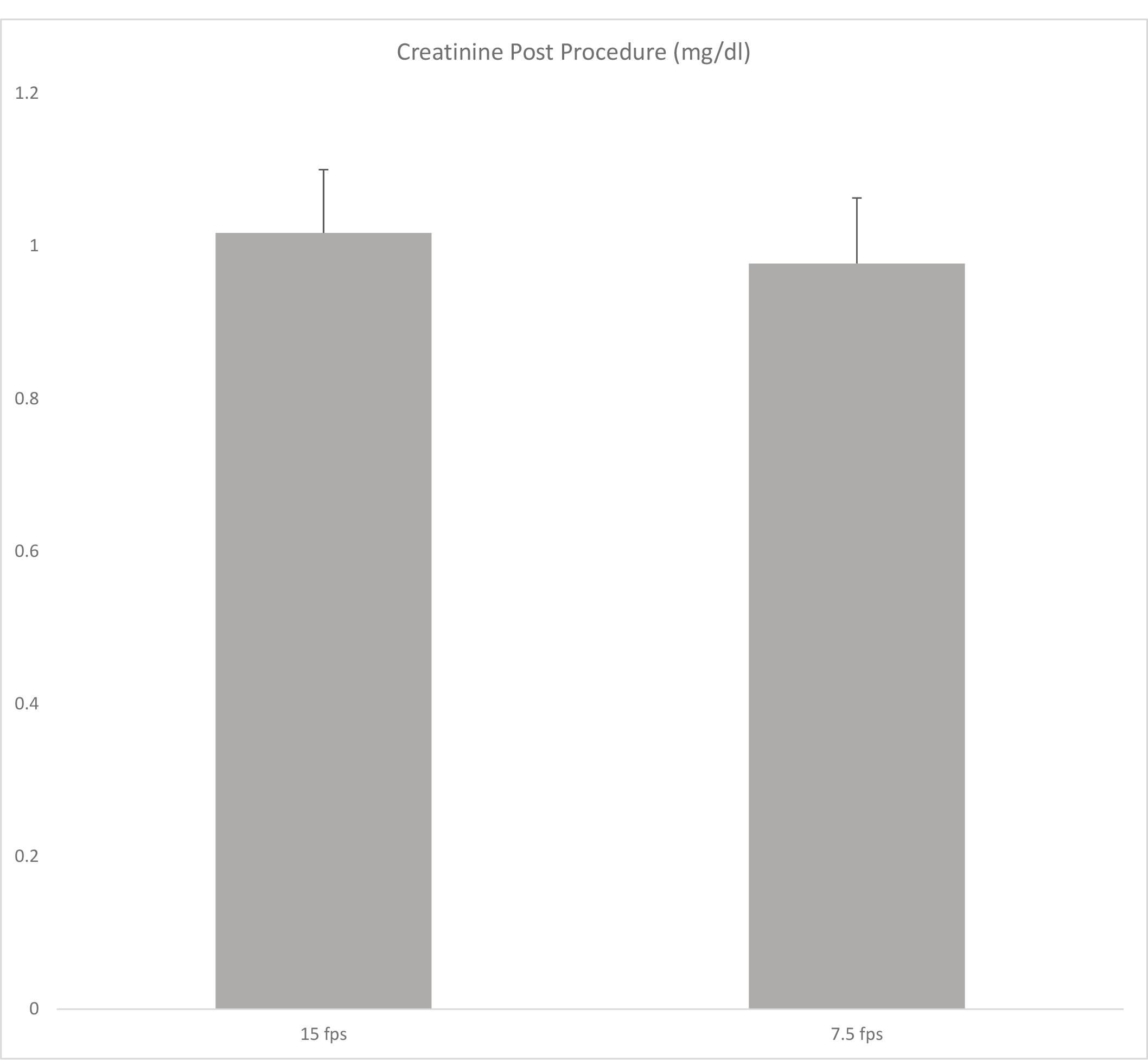
Creatinine post cardiac catheterization procedure amongst patients undergoing cardiac catheterization at 15-fps and 7.5-fps fluoroscopy rates

With respect to the primary outcomes, there were significant differences observed between the two group fluoroscopically. The cumulative air kerma for the 15-fps group was 524.140 mGy versus 266.576 mGy in the 7.5-fps group (p = 0.0018). Similarly, the total dose area product was also significantly lower in the 7.5 fps group (15335.617 mGy × cm^2^) compared to the 15-fps group (34784.095 mGY × cm^2^, p= 0.0003). Even with these differences in radiation, the total fluoroscopic time did not differ between the two groups (7.174 minutes in the 15-fps group and 8.462 minutes in the 7.5 fps group, p =0.4023). Furthermore, the change in renal function, which is a representation of the amount contrast utilized, did not differ between groups following catheterization. The creatinine 24 to 48 hours following catheterization was 1.017 milligram per deciliter (mg/dl) in the 15-fps group and 0.977 mg/dl in the 7.5 fps group (p =.7392).

The 30-day mortality rate was 0% in both the 7.5-fps group and the 15-fps. Similarly, there was no significant difference in the number of PCIs within 30 days between the two groups as only 6 patients in the 7.5-fps group and 3 in the 15-fps group required repeat PCI (Chi-Square P-value = 0.2899).

## Discussion

The present study aimed to evaluate the safety and efficacy of a frame rate of 7.5 fps during cardiac catheterization. The results showed that with a pulse a rate of 7.5 fps there was a significant radiation dose reduction compared to a rate of 15 fps. Patients in the 7.5 fps group had a significantly lower cumulative DAP (fluoroscopy and exposure), total DAP, and cumulative air kerma.

Importantly, there was no difference between the two pulse rates in terms of total fluoroscopic time. X-ray tubes produce pulses at rates that are at or below the video frame rate (usually 30 frames/second). When the X-ray pulse rate is lower than the video frame rate, the video frame corresponding to the last X-ray pulse is displayed repeatedly until the next X-ray pulse arrives. Traditionally, an X-ray pulse rate of 15 fps has been utilized to achieve a balance between video quality and radiation dose. Lower frame rates are associated with increased “jerkiness” and poorer image quality. In theory, this could compromise image quality and therefore lead to longer total fluoroscopic times. However, our results demonstrate that fluoroscopic time can be preserved even at a frame rate of 7.5 fps. This is important since fluoroscopic time is an important determinant of total patient radiation dose.^7^

These findings replicate results of other studies investigating the efficacy of a frame rate reduction from 15 fps to 7.5 fps in reducing radiation dose.^8-10^ In one retrospective cohort study, a conventional PCI protocol with 15 fps was compared with a protocol using 7.5 fps during fluoroscopic guidance and 10 fps during cineangiographic acquisition as well as selective fluoroscopic image storage.^8^ That study found a significant reduction in total air kerma and total DAP in the radiation reduction protocol compared with the conventional protocol. Moreover, total fluoroscopic time were similar between the two groups. Our study demonstrates that a frame rate of 7.5 fps can be maintained even during acquisition with a similar effect.

Furthermore, we showed that a frame rate of 7.5 fps did not lead to inferior clinical outcomes. There was no increase in the incidence of acute kidney injury from 15-to 7.5 fps. In addition, there was no difference in the thirty-day mortality between the two groups or in the rate of repeat PCI.

The study’s findings have important implications for clinical practice, as they suggest that reducing the frame rate during cardiac catheterization may reduce the risk of radiation exposure without compromising clinical outcomes. However, future studies are needed to confirm these findings and determine the optimal frame rate for cardiac catheterization procedures.

One limitation of this study is its relatively small sample size, which may limit the generalizability of the findings. Secondly, this was a retrospective study; therefore, there is attendant risk of selection bias. However, both groups did not differ in terms of background characteristics which should limit the impact of this bias. A strength of this study is that multiple metrics (DAP, air kerma) were used to measure radiation dose, which is an improvement from previous studies. Further research with a larger, prospective sample sizes and longer follow-up periods is needed to validate these findings and provide more robust evidence for clinical practice.

## Data Availability

The authors confirm that the data supporting the findings of this study are available within the article.

## Acknowledgements

None.

